# Human biting activity, resting behavior and yellow fever virus transmission potential of *Aedes* mosquitoes in southwest Ethiopia

**DOI:** 10.1101/2023.01.28.23285079

**Authors:** Abate Waldetensai, Myrthe Pareyn, Fekadu Massebo

## Abstract

**Background:** Yellow fever is an emerging and re-emerging viral disease transmitted through the bites of infective *Aedes* mosquitoes. Several outbreaks of yellow fever have been documented in southern Ethiopia.Understanding the transmission cycle is pivotal to manage arboviral disease outbreaks. Therefore, the present study aimed to evaluate which species of *Aedes* mosquitoes contribute to the YF virus transmission and the outbreaks that have occurred, and their behaviors (biting and resting) in the region.

**Methods:** Two districts were selected based on previous Yellow Fever (YF) outbreak history. A longitudinal entomological sampling was carried out to collect adult *Aedes* mosquitoes using human landing catches, mechanical mouth aspirators and pyrethrum spreadsheet collection. Adult mosquito collections were conducted twice a month for six months from February 2019 to July 2020. Identification of mosquito species at the genus level was done using morphological keys and speciation using molecular techniques. Aedes mosquitoes were pooled and tested for YFV, dengue virus (DENV, serotype 1-4) and chikungunya virus (CHKV) by qPCR.

**Principal findings:** A total of 1582 Aedes mosquitoes were collected; 669 (42.3%) from Boko Dawula and 913 (57.7%) from Ofa district. Of the 406 Aedes mosquitoes molecularly characterized to the species level, the *Aedes simpsoni* complex accounted for 99.5% (404/406), while *Aedes aegypti* found in the Ofa district accounted for only 0.5% (2/ 406). From the 934 *Aedes simpsoni* tested for viruses and none were positive. The human biting activities of *Aedes* (*Ae*.) *simpsoni* peaked at 8:00 – 9:00 hour and 16:00 – 17:00 hour, mostly outdoors, both within the villages and forests. The leaves of *Ensete (E*.*) ventricosum* appear to be ideal resting places for *Aedes (Ae*.*) simpson*i complex.

**Conclusion:** Although the tested *Ae. simpsoni* complex was negative for arboviruses; morning and afternoon activities of the species coincide with human outdoor activities and may therefore pose the risk of viral infection. The lower dominance of *Aedes aegypti* indicated that the major responsible vector for the occurrences of previous and current arboviral diseases was due to other mentioned Aedes species. It is of great importance to improve surveillance activities of arboviruses in reservoir hosts and vectors to establish control measures. Furthermore, the origin of bloodmeal and the mosquito’s role in the transmission of arboviral diseases need further study to improve the understanding of this species.

**Author Summary:** Aedes mosquitoes are the vector of most arboviruses infecting humans and animals. In Ethiopia, yellow fever (YF) outbreak is frequently occurring and claiming the lives of several people. Therefore, understanding the cycle of transmission is crucial in designing the prevention and control strategies for YF outbreaks. We conducted an entomological sampling in two districts with recent YF outbreaks in southwestern Ethiopia to identify and characterize the behavior and ecology of the Aedes mosquito species playing role in transmission. The *Ae. simpsoni* complex was the predominant species identified in both study areas and tends to bite humans in the morning and afternoon when most people are active outdoors within the villages and forests. Although none of the tested *Ae. simpsoni* complex mosquitoes were positive for arboviruses, improving surveillance activities in reservoir hosts, including primates and vectors, could be a key to establishing prevention and control strategies.

## Introduction

The emergence and re-emergence of arboviral diseases raise global concerns and threats to human health [1]. The viruses usually circulate among wild animals, but spill over to humans sporadically, leading to epidemics [2]. Arboviral diseases are increasingly causing morbidity in Ethiopia, with a significant increase in the number of outbreaks in the past years. Multiple outbreaks of yellow fever (YF) have occurred and affected the non-immune human population [3]. The first YF outbreak occurred between 1960 and 1962 when 100,000 cases and 30,000 deaths were reported in the southwest of the country [3]. The second YF outbreak took place between 2012 and 2013 in south Omo in southern Ethiopia [4]. As a response, 550,000 people were vaccinated in the region [5]. In 2016, 22 suspected YF cases and five deaths were reported in South Omo again [6]. In October 2018, another YF outbreak has been confirmed in Wolaita in southwest Ethiopia resulting in the death of 10 people [7]. Following confirmation of YF outbreaks, a vaccination campaign targeting 31,565 high-risk populations was conducted in October 2018 in six identified kebeles (villages; the lowest administrative unit). In addition, a ring vaccination campaign was carried out among 1,335,865 inhabitants in nine districts, seven in Wolaita zone and two in Gamo-Gofa zone [8]. Recently, the YF outbreak occurred in the Gurage zone in southwestern Ethiopia [9]. Although YF is endemic in the country, vaccination is not part of the childhood immunization program. Therefore, children and others living outside of vaccinated areas remain at risk for future YF outbreaks [6]. In addition to YF, outbreaks of other arboviral diseases such as dengue (DEN) and chikungunya (CHIK) are also common [10]. A previous study showed the presence of *Ae. simpsoni* complex in South Omo in villages where the 2012-2014 YFV outbreaks occurred [5].

Although arboviral disease outbreaks are common in Ethiopia, limited attention has been given to entomological monitoring and surveillance to identify appropriate vector control tools for outbreak prevention and control [10]. The vector mediating the periodic outbreaks is unknown and there is so far no evidence of whether the outbreaks are initiated by spillover from forest catchments into urban areas or other reasons [11]. Therefore, this study assessed the Aedes mosquito species composition, ecology, human biting activity and role in YFV transmission in sylvatic and human occupational spaces, in areas where YF outbreaks recently occurred in southern Ethiopia. This information is important to build on for further research eventually aiming to design appropriate vector control tools which might contribute to the prevention and control of future YF epidemics.

## Materials and Methods

### Ethical consideration

Ethical clearance was obtained from Arba Minch University Institutional Review Board (Ref. No IRB/108/11). Local administrative bodies and district health offices were informed and involved in the study. Written and oral consent was obtained from the study participants (households and data collectors). For entomological collections, YF-vaccinated volunteers were included and provided with anti-malarial prophylaxis.

### Study areas description

This study was conducted in Boko Dawula, former South Ari (South Omo zone) and Offa (Wolaita zone) districts in Southwestern Ethiopia (Figure 1). The two study districts were affected by YF outbreaks in 2016 (South Omo) and 2018 (Wolaita). The selection of the study districts was made in consultation with the zonal health office, village administrations and health extension workers. Both study sites are located on the edge of a forest and in both districts, the village closest to the forest was selected for mosquito collection.

**Figure 1:**
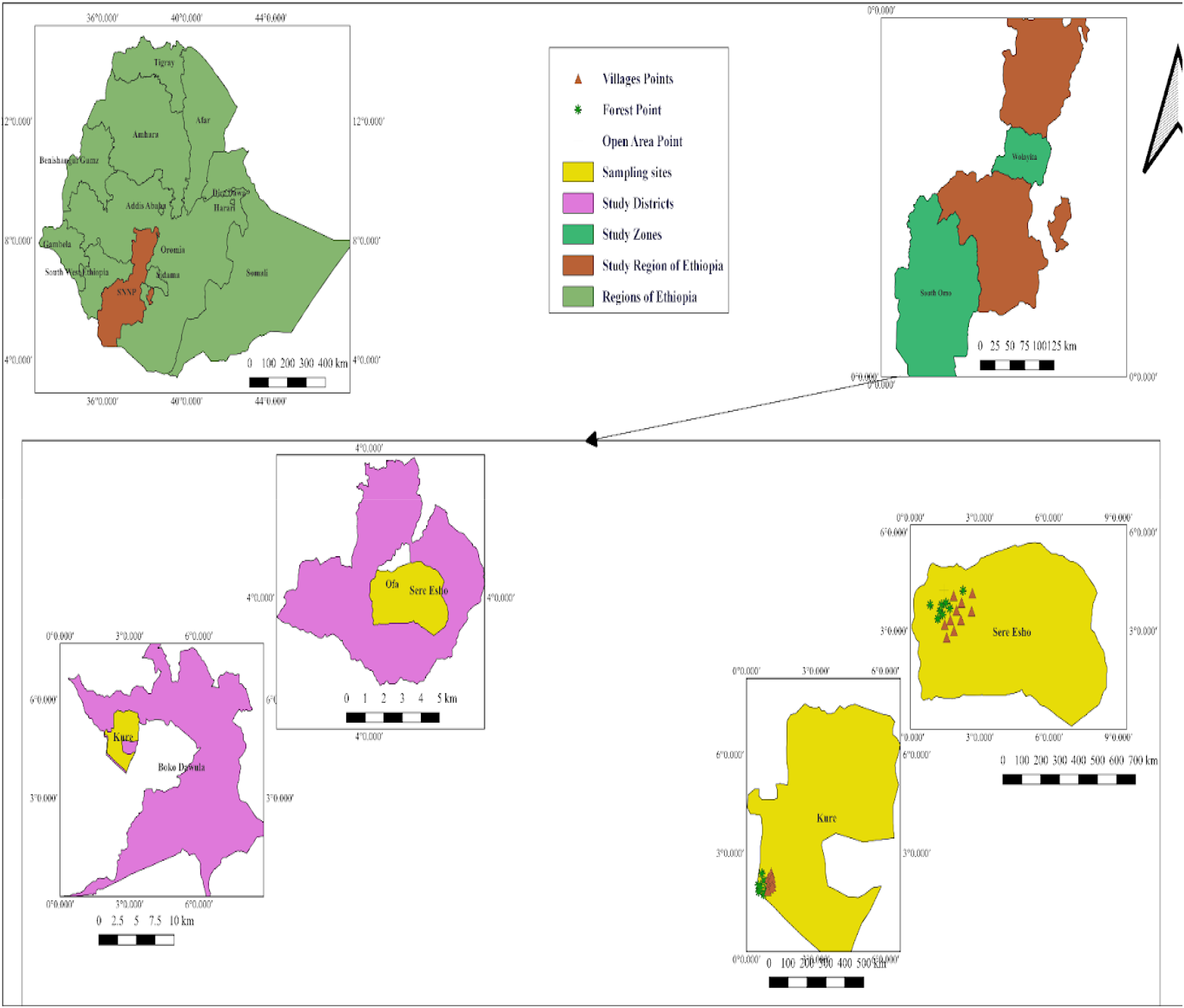
Map of the study sites in Boko Dawula district in the South Omo zone and Ofa district in the Wolaita zone, both in the Southern Nations Nationalities and Peoples’ Region (SNNPR) in southwestern Ethiopia.

Boko Dawula district is found at about 20 km south of Jinka town, the capital of the south Omo zone. The altitude ranges between 1000-1300 meters above sea level (masl) [12]. The mean annual rainfall ranges between 601-1600 mm and its mean annual temperature lies between 10-30°C [13]. Offa district is located 29 km southwest of the zonal capital, Wolaita Sodo. It is located at an altitude between 1000-2000 masl [14]. The amount of rainfall is between 800 and 1400mm with a temperature ranging between 14-28°C [15]. Due to high population pressure, agricultural expansion and deforestation are common. The edge of the forest is mainly used for agriculture. In forests and villages, there are natural and artificial breeding habitats for mosquitoes, such as Boko Dawula

### Study design and sampling techniques

We collected *Aedes* mosquitoes in three different habitats: forests (sylvatic cycle), agricultural lands located between human settlements and forests, and rural human settlements (villages) for six months from February 2019 to July 2020 taking into account both the Ethiopian dry (February - April) and wet months (May - July). Human landing catches (HLC), pyrethrum space spray collection (PSC), and mechanical aspiration (MAC) were employed to collect host-seeking and resting mosquitoes.

### Human landing collection (HLC)

Outdoor HLC was performed twice per month at three sampling habitats (villages, forest and agricultural lands) with a distance of at least 200m between sampling sites. In the forest and agricultural land, HLC was conducted at three sites in each district. In the villages of the two selected districts, HLC was conducted both indoors and outdoors in and around five selected houses. For each house, two collectors were assigned, one indoor and one outside with hourly rotation. The collection was performed between 6:00-10:00 and 15:00-20:00. Volunteers, wearing long-sleeve shirts were seated in chairs by exposing their legs below their knees to collect mosquitoes attempting to bite using an aspirator. Hourly collected mosquitoes were transferred into a paper cup and preserved as described below.

### Pyrethrum spray collection (PSC)

Indoor resting mosquito collection was done twice per month in fifteen randomly selected houses (other than HLC) in both villages, following the WHO indoor residual spraying (IRS) operational guideline [16]. The floor, beds and furniture were covered with white sheets, and food items were removed and properly closed before spraying an aerosol insecticide (Baygon, SC Johnson and Son Inc, Racine, Wisconsin, US) [17]. Additionally, windows, doors and other openings were closed to prevent mosquitoes from escaping. The collection was done early in the morning before smoking the houses. About 10 minutes post-spraying, mosquitoes were collected and preserved as described below [16].

### Mechanical aspirator collection

The MAC collection was performed twice per month at each site using a mechanical aspirator. Twelve houses were randomly selected (other than HLC and PSC) to collect mosquitoes resting on interior and exterior walls. For the collection of outdoor resting mosquitoes, four additional sites including caves, rock holes, tree holes and leaves were selected. A collector spent 10 minutes at each site searching for resting mosquitoes, using a timer. The number of mosquitoes collected and the type of resting site was recorded. To minimize collector bias due to skill variation, collectors were alternated between collection sites.

### Mosquito species identification

The collected mosquitoes were immediately identified at the genus level in the field using morphological keys [18,19]. *Aedes* mosquitoes were preserved individually in RNA later, transported in a coldchain and stored as soon as possible at -20°C for further analysis. From all collected *Aedes* mosquitoes, 406 were selected for molecular species identification taking into account the habitat, collection method, time and month; to make certain the representativeness of the sample. The legs of each specimen were used for DNA extraction using the NucleoSpin Tissue Kit (Macherey Nagel, Düren, Germany) according to the manufacturer’s instructions. Elution was done in 50 µL nuclease-free water. After that, the isolated DNA was subjected to a PCR targeting the mitochondrial *cytochrome oxidase 1* (COI) gene as described in [20]

Amplicons were checked on a 1.5% gel and sent for Sanger sequencing using the same primers. The DNA sequences were cleaned, and consensus sequences were generated and subjected to alignment using the ClustalW tool [21] implemented in MEGA X 10.1. Primers were trimmed to have sequences with equal lengths. Analyses of nucleotide composition and divergences among individuals were quantified using the Kimura 2-parameter (K2P) distance model [22]. A neighborhood joining (NJ) tree based on the K2P distances was made to provide a graphic representation of the clustering pattern among different species [23]. The presence or absence of a barcode gap was also determined for each species as a test of the reliability of its discrimination. This was to visualize and demonstrate the divergence in sequences obtained and to compare them to sequences held in GenBank. All sequences were compared with *Aedes* species sequences in the GenBank, GenBank accessions: [*Ae. simpsoni* (KT881398.1) and *Ae. aegypti* (MK300226.1)].

### Arboviral screening

For arboviral screening, mosquito specimens (head, thorax and abdomen) were pooled from 8 to 12 according to the habitat and month in which they were collected. Then, a 500 µL DNA/RNA shield (Zymo Research – Baseclear, Leiden, the Netherlands) was added to each mosquito pool and specimens were homogenized with a sterile pestle. Homogenates were centrifuged and the supernatant was transferred to a nuclease-free tube. RNA extraction was done using the Quick-DNA/RNA Pathogen Miniprep kit (Zymo Research - Baseclear, Leiden, the Netherlands) according to the manufacturer’s instructions, but the columns were replaced by Zymo-Spin IICR columns (Zymo Research). The RNA isolates were tested with three different assays: (i) a multiplex DENV 1-4 RT qPCR [20], (ii) a YFV RT-qPCR and, (iii) a CHIKV RT-qPCR [24] as described before. Commercial positive and negative (nuclease-free water) PCR controls were included in each PCR to validate the run.

### Data analysis

Data analysis was done in SPSS (version 21) software. Descriptive statistics (percentages, mean and standard deviations), were used to summarize the data. Generalized linear, model (ANOVA), chi-square, and t-test were used to compare vector density and behaviors of the vectors between villages/land cover types. Human biting rates (HBR) were measured directly from human-landing collections. In all analyses (*p* < 0.05) was considered significant. The hourly mean of the biting number of adult female mosquitoes was calculated using Pearson’s correlation.

## Results

### Mosquito species composition and ecology

A total of 1689 mosquitoes 93.7% (1582/1689) of *Aedes* and 6.3% (107/1689) of *Culex* were collected. Of the total collected mosquitoes, 58.7% (991) were from Ofa study sites whereas the left 41.3% (698) were from Boko Dawula. Most of the mosquitoes were collected during the wet season 97.9% (1653). Of the 1582 *Aedessimpsoni* complex, 54.1% (913/1582) were from Ofa district and the remaining 39.6% (669/1582) were from Boko Dawula district (Table 1).

**Table 1:** Mosquito species composition and seasonal distribution in Ofa and Boko Dawula districts, Southwestern Ethiopia

| Sites | Mosquitoes | Seasons |  |  |  |  |  |
| --- | --- | --- | --- | --- | --- | --- | --- |
|  |  | Dry |  | Wet |  | Total |  |
|  |  | N | Percent | N | Percent | n | Percent |
| Ofa | *Aedes* | 3 | 0.2 | 910 | 53.9 | 913 | 54.1 |
|  | *Culex* | 17 | 1.0 | 61 | 3.6 | 78 | 4.6 |
|  | Total | 20 | 1.2 | 971 | 57.5 | 991 | 58.7 |
| Boko Dawula | *Aedes* | 0 | 0.0 | 669 | 39.6 | 669 | 39.6 |
|  | *Culex* | 16 | 0.9 | 13 | 0.8 | 29 | 1.7 |
|  | Total | 16 | 0.9 | 682 | 40.4 | 698 | 41.3 |
| Total | *Aedes* | 3 | 0.02 | 1579 | 93.5 | 1582 | 93.7 |
|  | *Culex* | 33 | 2 | 74 | 4.4 | 107 | 6.3 |
| Grand Total |  | 36 | 2.1 | 1653 | 97.9 | 1689 | 100.0 |

A high number of *Aedes* mosquitoes was collected from tree leaves, E. *ventricosum* Caves and interior walls during wet seasons (June to July) (Figure 2).

**Figure 2:**
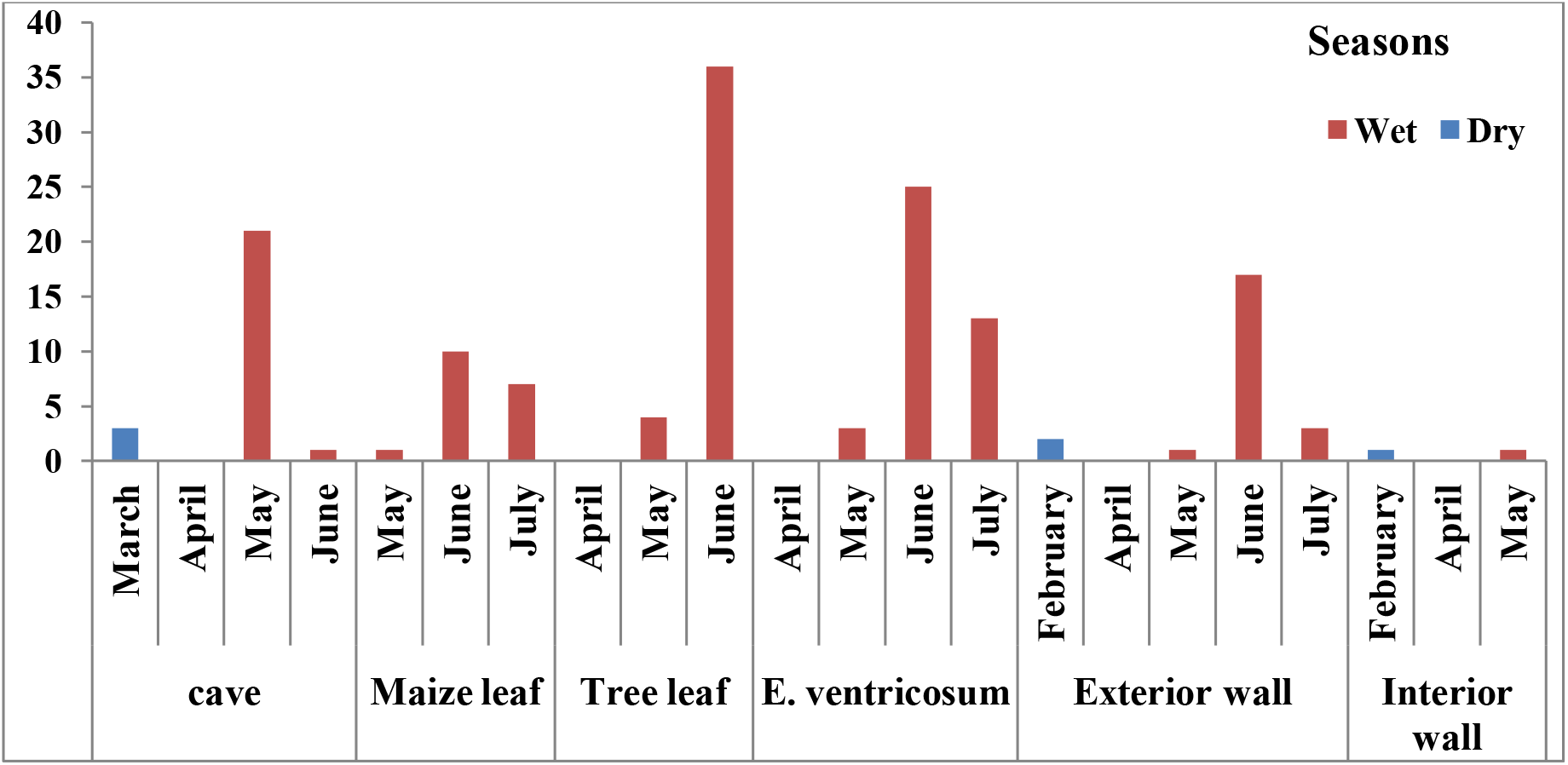
Ecological and seasonal distribution of Mosquitoes in Southwestern Ethiopia

The majority of *Ae. simpsoni* complex were collected by HLC (91.2%, n = 1442, mean = 15.02) from all land covers. Of these, 1475 (93.2%) were unfed whereas only 107 (6.8%) fed Aedes mosquitoes (Table 2).

**Table 2:** Abdominal condition of *Ae. simpsoni* complex from different collection sites by different entomological sampling techniques in southwestern Ethiopia

| Abdominal status | Land cover type | Methods |  |  | Total (mean, %) |
| --- | --- | --- | --- | --- | --- |
|  |  | Aspiration (mean, %) | HLC (mean, %) | PSC (mean, %) |  |
| Feed | Forest | 41 (3.4, 2.6) | 2 (0.2, 0.1) | 0 (-, 0) | 43 (3.6, 2.7) |
|  | Indoor | 1 (0.1, 0.1) | 0 (-, 0) | 0 (-, 0) | 1 (0.1, 0.1) |
|  | Outdoor | 63 (5.3, 4) | 0 (-, 0) | 0 (-, 0) | 63 (5.3, 4) |
|  | Total | 105 (2.2, 6.6) | 2 (0.5, 0.1) | 0 (-, 0) | 107 (2.23, 6.8) |
| Un feed | Forest | 30 (2.5, 2) | 451 (37.6, 28.5) | 0 (-, 0) | 481 (40.1, 30.4) |
|  | Indoor | 0 (-, 0) | 2 (0.2, 0.1) | 3 (0.25, 0.2) | 5 (0.4, 0.3) |
|  | Open area | 0 (-, 0) | 97 (8.1, 6.1) | 0 (-, 0) | 97 (8.1, 6.1) |
|  | Outdoor | 2 (0.2, 0.1) | 890 (74.2, 56.3) | 0 (-, 0) | 892 (74.3, 56.4) |
|  | Total | 32 (0.7, 2) | 1440 (30, 91) | 3 (0.1, 0.) | 1475 (30.7, 93.2) |
| Grand Total |  | 137 (1.43, 8.7) | 1442 (15.02, 91.2) | 3 (0.03, 0.2) | 1582 (16.5, 100) |

### Hourly biting activity

Of the total collected *Aedes* mosquitoes with HLC techniques (n = 1442) (Table 2), the hourly biting number of *Ae. simpsoni* complex significantly varied across collection times (F= 19.2; DF = 9; P ≤ 0.001). The highest peak biting time of *Ae. simpsoni* complex was observed between 16:00 and 17:00hour (32.5/hour/person). In the morning, the relative peak biting activity was reported between 8:00 and 9:00 hours (19.4/hour/person). The peak biting activity of *Ae. simpsoni* complex sharply declined after 10h in the morning and 18h in the afternoon (Figure 3).

**Figure 3:**
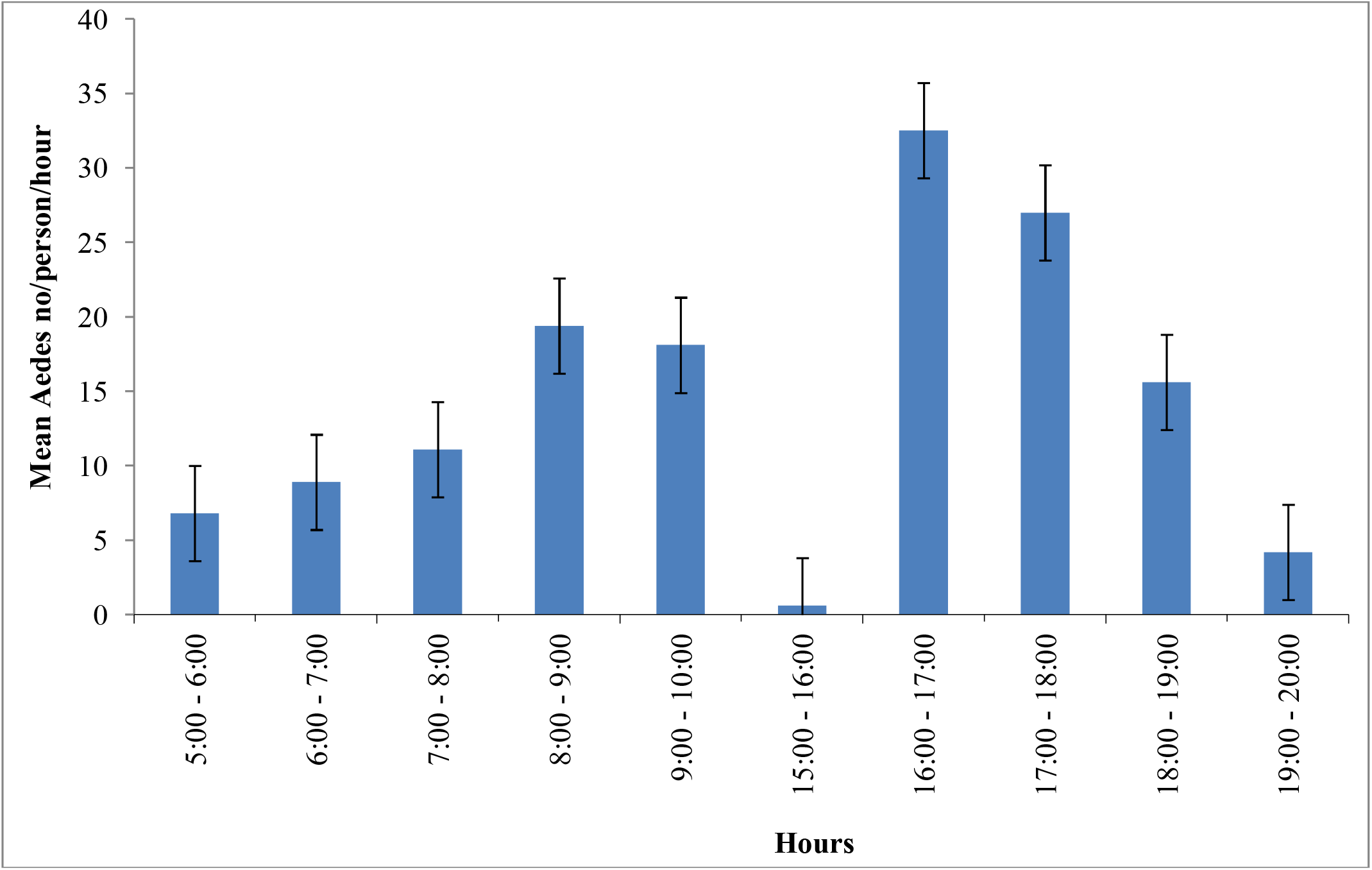
Overall hourly biting activity of *Ae. simpsoni* complex/person/hour in Boko Dawula and Offa districts, southwest Ethiopia

*Ae. simpsoni* complex was consistently active with a similar pattern outdoors within the villages and inside the forests. The peak human biting activity of *Ae. simpsoni* observed outdoors averaging 10.6 bites/person/hour in Offa and 6.3 bites/person/hour in Boko Dawula from 16:00-17:00hours. Morning peak human biting activity in the forest was observed from 8 to 10 am in both Ofa and Boko Dawula. The highest biting peak of *Ae. simpsoni* complex in the forest was observed between 16:00 and 17:00 hour (13.4/person/hour) in Boko Dawula. The peak biting times of Aedes was also relatively observed in Open land (Figure 4).

**Figure 4:**
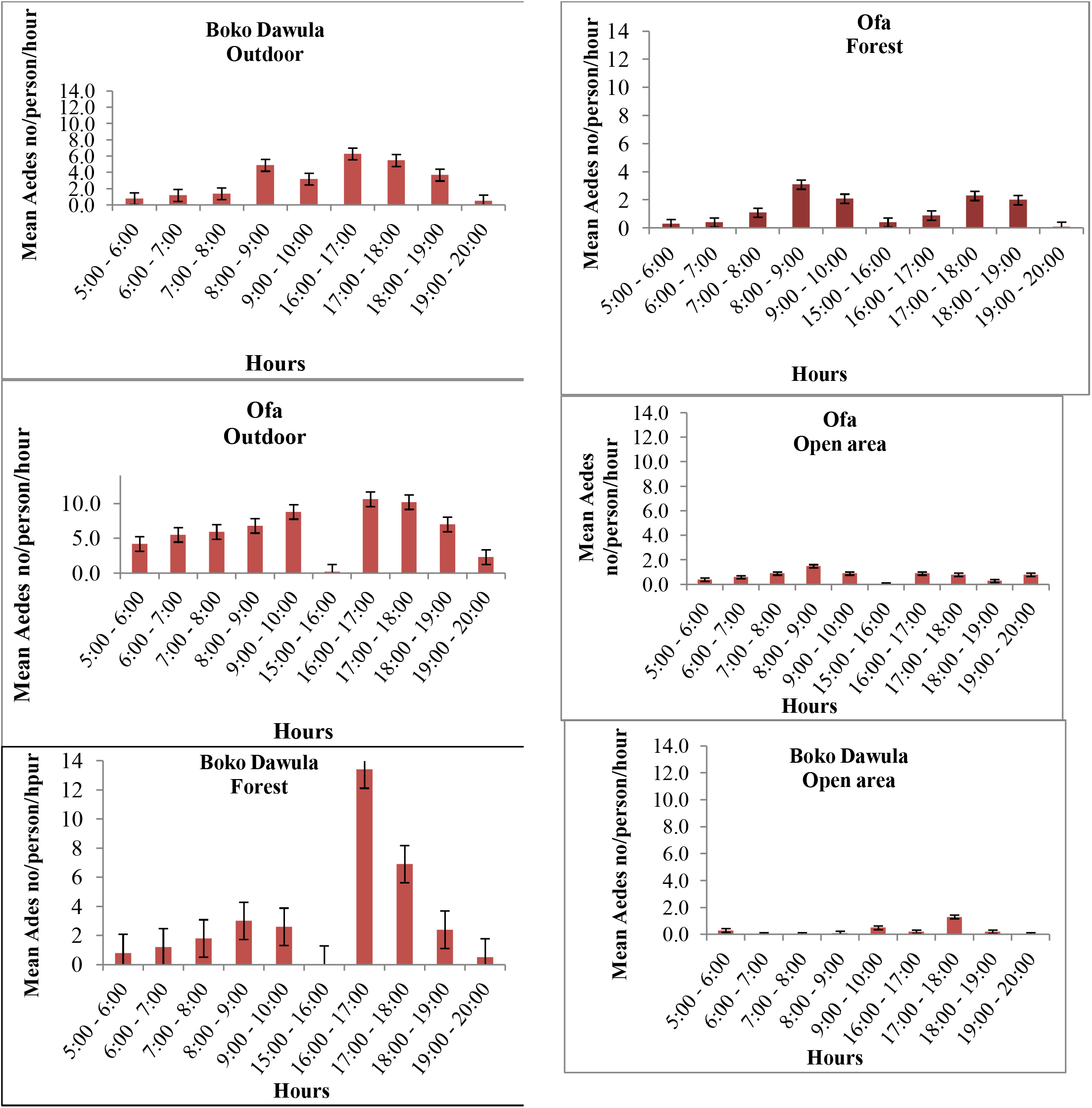
Hourly biting activity of Aedes simpsonicomplex/person/hour in Boko Dawula and Offadistricts, southwest Ethiopia

**Figure 5:**
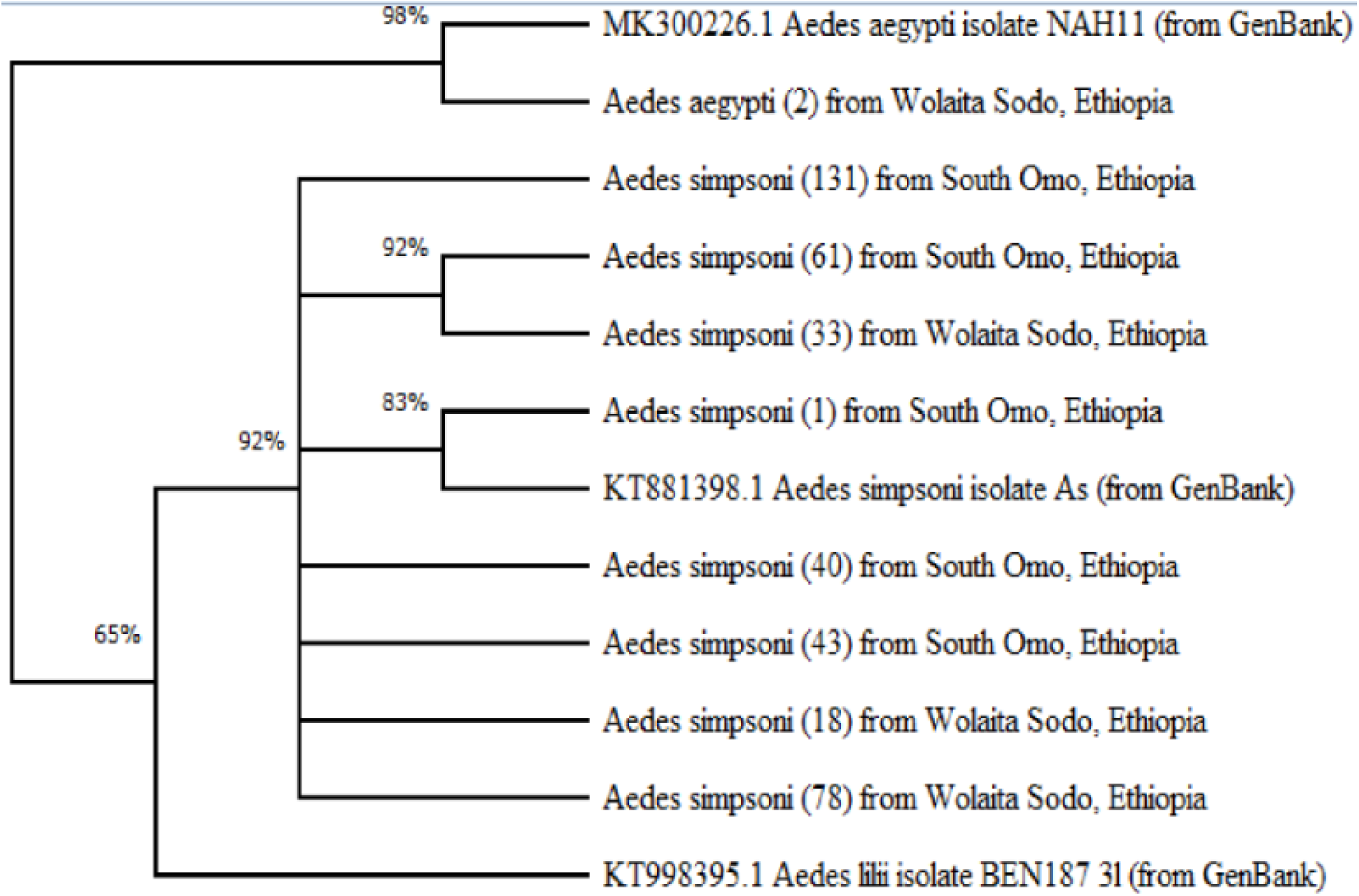
Evolutionary analysis by evolutionary likelihood method and Tamura-Nei model: a phylogenic tree

### Resting sites

To assess resting behavior, *Ae. simpsoni* complex collected by MAC was considered, by which 137 Aedes mosquitoes were collected overall. The density of the *Ae. simpsoni* complex varied by resting sites (*P*< 0.001, Df = 11). Relatively larger numbers of *Ae. simpsoni* were collected from *E. ventricosum* followed by tree leaves (Table 3). A single *Ae. simpsoni* complex was collected on the interior wall. Overall, most *Ae. simpsoni* were collected during the rainy months of May, June and July.

**Table 3:** Densities of Aedes simpsoni complex from resting sites by mouth aspirators in southwest Ethiopia

| Collection sites | February | March | April | May | June | July | Total | Median | IQR |
| --- | --- | --- | --- | --- | --- | --- | --- | --- | --- |
| Cave | 0 | 3 | 0 | 15 | 1 | 0 | 19 | 0.5 | 2.5 |
| *E. ventricosum* | 0 | 0 | 0 | 3 | 25 | 13 | 41 | 1.5 | 10.5 |
| Maize leaf | 0 | 0 | 0 | 0 | 10 | 7 | 17 | 0 | 5.25 |
| Leaves | 0 | 0 | 0 | 3 | 35 | 0 | 38 | 0 | 2.25 |
| Interior wall | 0 | 0 | 0 | 1 | 0 | 0 | 1 | 0 | 0 |
| Exterior wall | 0 | 0 | 0 | 1 | 17 | 3 | 21 | 0.5 | 2.5 |
| Total | 0 | 3 | 0 | 23 | 88 | 23 | 137 | 13 | 22.25 |
| Total median | 0 | 0 | 0 | 2 | 13.5 | 1.5 | 20 | 0.75 | 1.875 |
| IQR | 0 | 0 | 0 | 2 | 19.75 | 6 | 16.25 | 1 | 5 |

IQR \*\*Inter-quartile Range

### Molecular speciation of *Aedes* mosquitoes

Of the 406 *Aedes* (198; 48. 8% from South Ari and 208; 51.2% from Ofa) mosquitoes screened for species identification, 404 (99.5%) were *Ae. simpsoni* complex. The other 404 *Aedes* mosquitoes had more than 84% closeness to the *Ae. simpsoni*. Two Aedes mosquitoes collected in the Ofa district had 99% proximity to Ae. aegypti in the GenBank.

### Screening for viral infection

Of the 934 Ae. simpsoni complex screened for viral infection by RT-qPCR, none of the pools was positive for YFV, DENV, serotype 1-4, or CHKV.

## Discussion

The current study was conducted to assess the *Aedes* mosquito species composition, ecology, human biting activity and their role in viral transmission in selected districts with a previous history of YF outbreaks in southwestern Ethiopia. *Ae. simpsoni* complex was the dominant *Aedes* mosquito species in the two districts. In the current study sites, there is no previously conducted biting behavioral studies on *Aedes* mosquitoes. The species was predominantly captured outdoors, particularly for human biting activity. Majority of the *Aedes* mosquito were collected outdoors and within the forest.Very rarely, Aedes mosquitoes were collected indoors.This indicated that *Aedes* mosquitoes are commonly biting human outdoors and within the forests. The current observation supports the finding of Captain-esoah et al.(2020) who reported larger numbers of *Aedes* biting outdoors than indoors [25]. In consistent to the study conducted in southeastern Senegal by [26], the outdoor biting behavior of *Aedes* might lead to an increase in risk of exposure toYF transmission outdoors than indoors.

*Aedes simpsoni* complex specifically *Ae. bromeliae* and *Ae. aegypti* have both previously been documented as the responsible vectors for the reemergence of yellow fever outbreak in South Ari, southwest Ethiopia [3]. Adult mosquitoes have been collected both indoors and outdoors in south Omo (South Ari) and were identified as *Ae. simpsoni* complex [5]. *Ae. simpsoni* complex was the predominant species in these previous studies; which may imply that the species is adapted to the sylvatic environment. in contrast to our study, in the previous tudy of [5], adult mosquitoes collected from south Omo were most likely *Ae. lilii*. In contrast, *Ae. aegypti* was dominant in Dire Dawa town, an urban setting in the eastern part of the country where YF outbreaks previously occurred [11]. This might be due to differences in the sylvatic and urban environments could be different and the species distribution could vary due to the adaptabilityor the species’ ability to adapt to particular environments. The same study screened *Ae. simpsoni* complex for arboviral infection and none of them were positive for YFV and other arboviruses.

Majority of the host seeking *Ae. simpsoni* complex mosquitoes were collected outdoors within the village and forest. The tendency of *Ae. simpsoni* complex to bite humans inside the forest and outdoors within the village might increase human exposure to YFV infection when they go into the forests to cut trees for agricultural expansion and other activities. Moreover, this behavior of *Ae. simpsoni* complex might make them suitable as a vector for the transmission of YFV in the sylvatic cycle.

The human biting activity of *Ae. simpsoni* was found to be high in the afternoon, with peak biting time at 16:00-17:00h both within the villages and forests. Peak biting times are usually altered to times when their potential hosts are less protected [25]. This indicates the highest risk to get into contact with *Ae. Simpsoni complex* is during outdoor activities in the afternoon. However, the mosquitoes are also quite active in the morning, when lots of human activities take place in the forests as it is generally colder then. This might increase the risk of infection due to high-human vector contacts [27,28].

*Aedes simpsoni* complex preferred to rest outdoors under the leaves of *E. vetricosum* and other plants. *E. vetricosum* was previously reported as the main breeding habitat for the same species in Southern Ethiopia [3,5]. This plant has a stem resistant to evaporation and stores water for a long time which makes it ideal for mosquito breeding and resting [29]. This implies that the *E. vetricosum* could be a potential site for monitoring and surveillance of both immature and adult stages.

None of the *Ae. simpsoni* complex screened for arboviral diseases was found to be positive. A few specimens of the same species from south Omo (South Ari) have been screened for different arboviruses previously and none of the were also positive [5]. The absence of virus circulation in this species might be due to the small number of mosquitoes screened or the absence of the movement of viremic vertebrates. Moreover, the YF virus transmission depends on the density of mosquitoes and wild primates [30]. During the outbreak of YF in the two districts, the majority of the community was vaccinated, which might also contribute to the absence of the virus in the vector population [31].

Additional to *Ae. Simpsoni* complex, our study also found 2 *Ae. Aegypti* mosquitoes in village habitats during afternoon times with outdoor HLC method. In contrast to the current study, high abundance of *Ae*.*aegypti* was reported in Dire Dawa, Eastern Ethiopia by [10] from immature stage collection. This indicates that *Ae. aegypti* complex is exophagic in parallel to the study conducted in Northern Ghana, that confirmed the *Ae. aegypti* outdoor biting tendency [25]. Similarly, the study from southeastern Senegal documented the outdoor biting tendency of *Ae. aegypti* [26]. Similar reports in Northern Ghana was shown that adult *Aedes spp*. are diurnal feeders, with exophilic feeding behavior during the early or late hours of the day, usually bite and rest outdoors before and after feeding [25].

Despite regular outbreaks of YF in Ethiopia, very little is known about the vectors and their behavior. However, this information is pivotal to designing vector control tools to prevent and control future YF epidemics. Our research findings give new insight into the temporal and spatial dynamics of the unusually *Aedes mosquito* species as a vecors of arboviral diseases transmission and its amplifications in their animal reservoirs that result in spillover infection of humans in an area. While many vectors may participate in maintenance of sylvatic Arboviral diseasas, Ae. simpsoni complex is most likely to be responsible for spillover into humans due to its broad land cover preferences and rates of human contact within village perimeters. This information can be used to inform the local population of the places and times of greatest risk for exposure so that mosquito avoidance or protective measures can be implemented.

### Conclusions

*Aedes simpsoni* complex was the most dominant species in the region and could be a potential vector responsible for the transmission of YFV. The human biting activity of *Ae. simpsoni* complex was predominantly in the morning (8:00 – 9:00hr) and afternoon (16:00 – 17:00hr) both outdoors within the villages and inside the forests, where people are active. Screening more mosquito samples are recommended to identify the most important vectors mediating transmissions. Future research should focus on sylvatic vector distribution at geographic large areas, blood meal sources, including reservoirs, using diversified traps types to collect *Aedes* before, during the outbreak and after the outbreak. Assessing the blood meal sources for a comprehensive assessment of arbovirus risk and estimating the reservoir hosts of YFV should be required.

## Data Availability

The data can be available with the corresponding author and provided up on request

## Data Availability

The data can be available with the corresponding author and provided up on request

## Acknowledgments

We are very grateful to South Omo and Wolaita zone Health department office leaders, the village head, Health extension workers, and volunteers for their cooperation during data collection. We are so proud to express our deep gratitude to the data collectors for their strong commitment to collecting mosquitoes. Our special thanks go out to Girum Tamiru (Arba Minch University) and Natalie van Houtte of the Evolutionary ecology group (University of Antwerp, Belgium) and Joachim Mariën from the Outbreak Research Team (Institute of Tropical Medicine Antwerp, Belgium) for their technical assistance in the laboratory process and activities.

## Author contributions

**Abate Waldetensai**:

Conceptualization, Methodology, Resources, Investigation, Visualization, Validation, Data curation, Formal analysis, Writing-original draft, and Writing-review and editing

**Fekadu Massebo:**

Conceptualization, Methodology, Resources, Supervision, Project administration, Visualization, Validation, Formal analysis and Writing-review and editing,

**Myrthe Pareyn:**

Methodology, Supervision, Visualization, Validation, Formal analysis, Writing-original draft and Writing-review and editing

## Data availability

All relevant data are within the manuscript are available at https://doi.org/10.6084/m9.figshare.19188638.https://doi.org/10.6084/m9.figshare.19188641.https://doi.org/10.6084/m9.figshare.19188644, https://doi.org/10.6084/m9.figshare.19188647

## Supplimentary information

Supplemental information for this article can be found online at https://doi.org/10.6084/m9.figshare.19187822.https://doi.org/10.6084/m9.figshare.19187843.https://doi.org/10.6084/m9.figshare.19187849.https://doi.org/10.6084/m9.figshare.21960107.https://doi.org/10.6084/m9.figshare.19187660.https://doi.org/10.6084/m9.figshare.19187657.https://doi.org/10.6084/m9.figshare.19187654.https://doi.org/10.6084/m9.figshare.19187642

